# Investigating cerebral anomalies in preterm infants and associated risk factors with MRI at term-equivalent age

**DOI:** 10.1101/2025.06.23.25330099

**Authors:** Nicolas Elbaz, Valérie Biran, Chloé Ghozland, Laurie Devisscher, Aline Gonzales-Carpinteiro, Aurélie Bourmaud, Lucie Hertz-Pannier, Yann Leprince, Alice Frérot, Alice Héneau, Jessica Dubois, Marianne Alison

**Author notes:** **Corresponding Author:** Jessica Dubois. co-last authors (Jessica Dubois, Marianne Alison).

## Abstract

**Background:** Being born very or extreme preterm is a major source of cerebral anomalies and neurodevelopmental disorders, whose occurrence depends on many perinatal factors. A better understanding of these factors could be provided by cerebral Magnetic Resonance Imaging (MRI) at term-equivalent age (TEA).

**Objective:** To investigate, through cerebral TEA-MRIs, the relationship between the main perinatal factors and the occurrence of cerebral anomalies, and cerebral volumetry.

**Methods:** We assembled a cohort of very and extremely preterm babies who underwent a cerebral TEA-MRI. We assessed cerebral anomalies using a radiological scoring system – the Kidokoro scoring – and performed cerebral volumetry. We investigated the relationship between 9 clinical factors (birth characteristics, resuscitation treatments…), volumetry and Kidokoro scores, and the relationship between Kidokoro scores and volumetry.

**Results:** Among 110 preterms who underwent a cerebral MRI at TEA, 7% suffered moderate-to-severe brain anomalies. We identified mechanical ventilation as a risk factor for cerebral anomalies (adjusted Odds-ratio aOR = 4.6, 95% Confidence Interval CI [1.7-12.8]) and prolonged parenteral nutrition as a protective factor (aOR = 0.2, 95%CI [0.1-0.7]) for white matter anomalies. Mechanical ventilation (p = 0.009) and being born small for gestational age (p < 0.001) were risk factors for the reduction of cerebral volumes. An increase in brain lesion severity was associated with decreased cerebral volumes (p = 0.017).

**Conclusion:** Our study highlights the importance of treatment-related perinatal factors on the occurrence of cerebral anomalies in very and extreme preterms, and the interest in using both qualitative (Kidokoro scoring) and quantitative (volumetry) MRI-tools.

## Introduction

Prematurity is a frequent phenomenon, occurring in approximately 10% of all births worldwide [1]. It is one of the main cause for neurodevelopmental disorders (NDD), especially in very and extreme preterm births (occurring before 32 weeks of gestation) (2% of all births) [2] [3]. Those disorders imply different kinds of limitations [4], among which cerebral palsy [5] or autistic spectrum disorders [6], and therefore lead to different degrees of disability. Since NDDs often arise from prematurity-associated brain anomalies [7], it was stated that neuroimaging through Magnetic Resonance Imaging (MRI) at term-equivalent-age (TEA) may help predict the neurodevelopmental prognosis of the premature child [8]. Indeed, brain MRI provides a large number of analytic tools, either qualitative or quantitative, based on both routine and advanced imaging techniques [9]. Such insights on the neurodevelopmental prognosis of preterms could allow for better risk stratification and optimized care strategies.

Additionally, the occurrence of brain anomalies in preterms and the ensuing disorders are themselves influenced by many perinatal clinical factors, that mainly represent risk factors [10]. Among them, pro-inflammatory factors play a major role. For example, prenatal chorioamnionitis can lead to reduced cerebral volumes [11] and later cognitive performances in children [12]. Postnatal necrotizing enterocolitis (NEC) has been associated with white matter lesions [13] and cerebral palsy [14]. A thorough knowledge of those clinical factors is thus essential to evaluate the prognosis of preterms. Yet, the role of those factors in brain resilience or vulnerability is not fully understood.

To better understand the clinical factors affecting brain development in preterms during the perinatal period, we aimed at investigating the relationship between the main early clinical factors and the occurrence of cerebral anomalies in the very and extremely preterm infants: using MRI-scans at TEA, we searched first for an association between clinical factors and a global brain abnormality score developed for preterms by Kidokoro et al. (or “Kidokoro scoring”) [15] and widely used by clinicians, and secondarily for an association between the same factors and measures of brain volumetry.

## Methods

### Population and study design

This is a monocentric cohort study of neonates admitted between April 2021 and July 2024 to tertiary neonatal intensive care units (NICU) right after birth.

Preterm neonates were born in two different maternities and TEA-MRI was performed on the same 3T MRI scanner with a standardized protocol. We included all preterm neonates born at a gestational age below 32 weeks, that had undergone a TEA-MRI between 38 to 43 weeks of post-menstrual age (PMA). The requirements to undergo a TEA-MRI were either to be born extremely preterm (before 28 weeks), or to be born very preterm (between 28 to 32 weeks) with cerebral anomalies on cranial ultrasonography. We excluded the neonates with significant malformations or genetic/syndromic disorders.

This study was approved by the Ethic Committee of research in medical imaging, of the CERF (Collège des Enseignants en Radiologie de France), and the CPP (Comité de Protection des Personnes) Ile de France 3 (protocol DEVine, CEA 100 054). All procedures were carried out in accordance with the ethical principles established in the Declaration of Helsinki and local regulations regarding data protection.

### TEA-MRI

TEA-MRI was conducted on an Ingenia Philips Healthcare 3.0 Tesla MR system, using a multi-channel head coil, without sedation (spontaneous sleep was obtained after feeding).

MRI protocol consisted of a systematic multimodal evaluation, including:

- 2D T2-weighted imaging in coronal, axial and sagittal planes (echo time [TE] = 150 ms, repetition time [TR] = 5500 ms ; slice thickness = 2.0 mm, flip angle 90° ; FOV = 123 x 140 x 153 mm ; matrix 192 x 192 ; reconstructed voxel size 0.8 x 0.8 mm^2^), total duration 3’18’’, 3’18’’ and 3’51’’
- A convention sagittal 3D T1-weighted imaging (TE = 33 ms, TR = 500 ms ; flip angle 90° ; FOV = 170 x 176 x 140 mm ; matrix 224 x 224 x 175 ; resolution 0.79 x 0.79 x 0.8 mm), duration 3’54’’
- An axial 3D Susceptibility Weighted Imaging SWI (TE = 6 ms, TR = 40 ms ; flip angle 17° ; FOV = 157 x 179 x 140 mm ; resolution 0.8 x 0.8 x 1.5 mm), 4’13’’
- An axial diffusion-weighted sequence (with a b value of 0 and 1000 s/mm^2^ for six directions of diffusion gradients) for computation of Apparent Diffusion Coefficient (ADC) (TE = 75 ms, TR = 5500 ms ; FOV = 160 x 160 x 108 mm ; matrix 80 x 80 x 54 ; 2-mm isotropic resolution), duration 2’04’’.

### Qualitative analysis of MRI: Kidokoro scoring

Analysis of MRI was performed on a workstation (Carestream Vue PACS software) by two operators: a senior and a junior radiologist. In case of disagreement, a consensus was obtained (MA and NE).

Qualitative analysis was performed by the same operator (junior radiologist NE) using standardized radiological Kidokoro scoring system [15]. An abnormality score and degree of severity were thus obtained for each cerebral compartment: grey matter GM (cortex and central grey nuclei), white matter WM, cerebellum, and for the whole brain. Cortical abnormality was graded based on 3 variables: signal abnormality, delayed gyration and increased extracerebral space; while central grey nuclei abnormality was graded based on signal abnormality and gross volume reduction (evaluated by the deep GM area). GM score was classified as normal (0), mild (1), moderate (2), or severe (≥ 3).

The occurrence of cerebral WM injury was graded based on 6 variables: cystic lesion, focal signal abnormalities, delayed myelination, thinning of the corpus callosum, dilated lateral ventricles and gross reduction of WM volume (evaluated by the biparietal diameter). WM score was classified as normal (0-2), mild (3-4), moderate (5-6), or severe (≥ 7).

Cerebellum also was graded based on signal abnormality and gross volume reduction (evaluated by transverse cerebellar diameter). Cerebellum score was classified as normal (0), mild (1), moderate (2), and severe (≥ 3).

Finaly, a whole brain score was calculated as a sum of the three regional subscores and classified as normal (0-3), mild (4-7), moderate (8-11), and severe (≥ 12).

Presence of hemorrhagic lesions was analyzed according to Papile classification. Diffusion weighted imaging was further used to perform quantitative ADC measurement in right and left frontal and parietal WM. An increased ADC value according to normative values (beyond 1.7 mm^2^/s) [16] was classified as “Diffuse excessive high signal intensity (DEHSI)”.

For the Kidokoro scoring, a reproducibility study was performed on twenty subjects. These subjects were selected according to their degree of severity, in order to obtain a fair distribution of each category. A second evaluation was performed by the same operator at a distance from the first assessment, to estimate an intra-observer concordance coefficient.

### Quantitative analysis: cerebral volumetry

Besides, cerebral volumetry was performed using state-of-the-art tools. We first reconstructed a T2-weighted volume with a 0.8 mm isotropic resolution, using the super-resolution tool NiftyMIC [17] based on at least two different orientations (coronal, axial or sagittal) without movement artefacts. The resulting images were used to segment the different brain tissues and compartments by coupling two automatic tools dedicated to the neonatal brain: DrawEM (Developing brain Region Annotation With Expectation-Maximization) [18] and iBEAT (infant Brain Extraction and Analysis Toolbox) [19]. As each segmentation showed its own qualities and shortcomings, they were combined (by masking and applying tools of mathematical morphology) to obtain the following compartments in the most reliable and reproducible way possible: cortical grey matter, central grey matter, sub-cortical and central white matter (combined), cerebellum, brainstem, hippocampus and cerebro-spinal fluid (**Figure 1a**). Visual inspections by the same operator (NE) highlighted some significant errors and inaccuracies in about 25% of segmentations: each segmentation was assessed and received a grading: “perfect”, “good”, “acceptable”, “bad” and “unusable”. The “bad” and “unusable” segmentations had to undergo manual corrections (by AGC and JD) and further inspection (by NE). The compartment volumes were then quantified.

**Figure 1:**
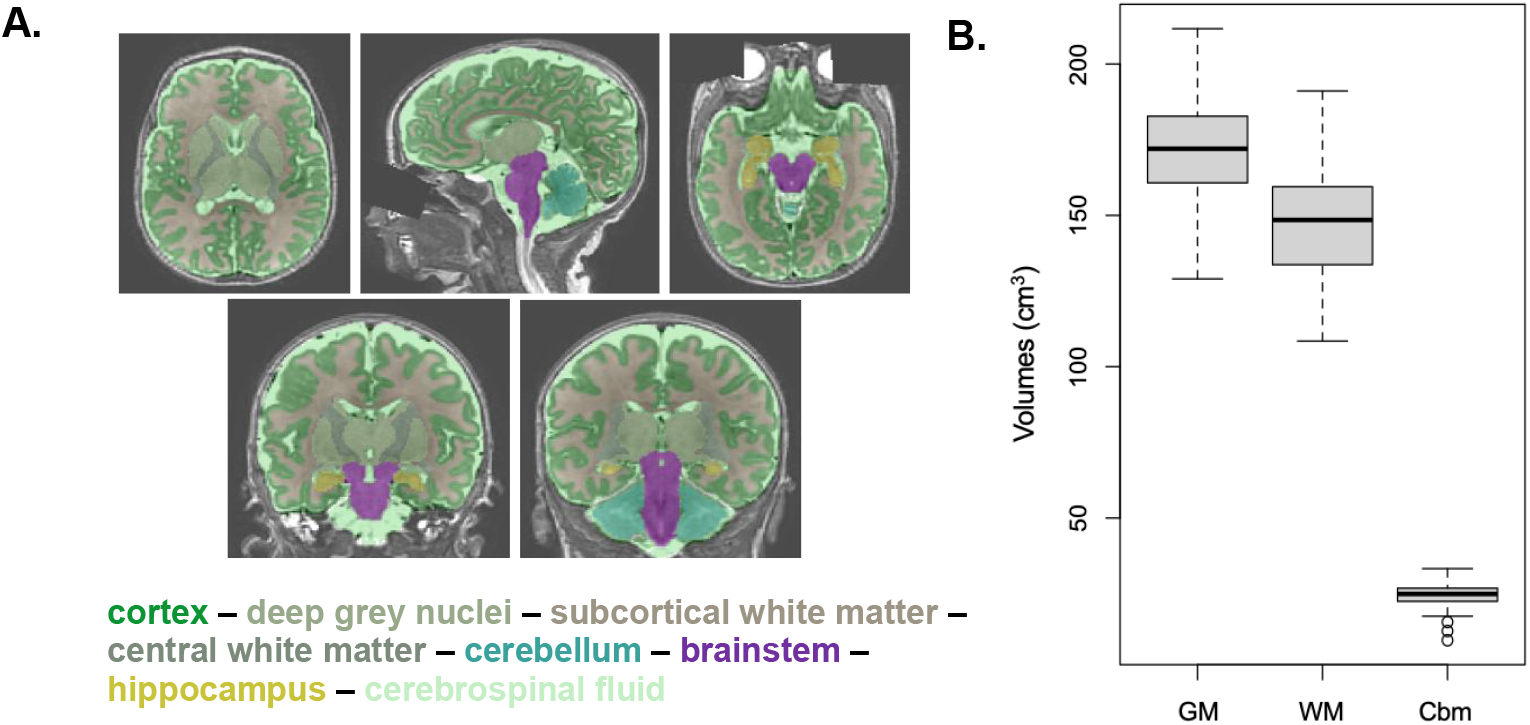
**A**. Segmentation of cerebral compartments according to a combination of iBEAT and DrawEM methods: cortex in emerald green, deep grey nuclei in sage green, subcortical white matter in white, central white matter (internal capsule) in grey, cerebellum in blue, brainstem in purple, hippocampus in yellow, cerebrospinal fluid in mint green. **B**. Box-Plot describing cerebral volumetry over the group of infants for the different brain compartments: grey matter GM (cortex and deep grey nuclei), white matter WM (subcortical and central) and cerebellum Cbm.

### Clinical characteristics

For each patient, clinical data referring to birth (gestational age at birth, birth weight, sex) and the presence of severe neonatal morbidities (such as necrotizing enterocolitis, and bronchopulmonary dysplasia at 36 weeks of postmenstrual age) were prospectively recorded into the medical chart.

Clinical data of interest were then collected retrospectively using the hospital electronic health record system. We inquired for 9 clinical factors, considered to be among the main neonatal morbidity risk factors: category of gestational age at birth, sex, monochorionic twin pregnancy, being small for gestational age at birth (SGA), perinatal sepsis, necrotizing enterocolitis (NEC), bronchopulmonary dysplasia (BPD), use of prolonged parenteral nutrition, and use of mechanical invasive ventilation.

The sex of the preterm was considered as moderate-to-severe NDDs are more frequent in preterm boys, hinting for a protective role of female sex [20]. We considered specifically monochorionic twin pregnancies as the occurrence of brain lesions is higher compared with dichorionic ones, due to vascular complications [21]. Gestational age was estimated based on first trimester ultrasound, and three groups were considered: born below 28 weeks PMA (G1), between 28 to 30 weeks (G2), and between 30 to 32 weeks (G3). Infants were classified as small for gestational age if they were born with a birth weight below the 10^th^ percentile on customized AUDIPOG curves for male and female neonates [21]. Sepsis was defined as any infection requiring antibiotic treatment for longer than five days, whether a bacterial pathogen had been detected or not. NEC diagnosis was based on both clinical data and imaging according to Bell’s classification [22], only stage ≥ II NEC were considered. BPD was defined as a requirement for oxygen-therapy beyond 36 weeks of PMA or for more than 28 days after birth. Prolonged parenteral nutrition was defined as a continued parenteral nutrition for more than 21 days. Mechanical invasive ventilation was only considered if it lasted strictly more than one day.

### Statistical analysis

Data were analysed using the R 4.3.3 programming software (R Core Team 2025). Discrete and categorical variables (the 9 clinical factors, Kidokoro score and injury severity according to Kidokoro scoring, hemorrhagic lesions, DEHSI) were expressed as frequency and percentage. Continuous variables (PMA at TEA-MRI, cerebral volumes) were expressed as mean and standard deviation as appropriate. Hemorrhagic lesions and DEHSI were reported here for descriptive purposes only, and were not considered for regression analysis.

The relationship between Kidokoro injury severity and the nine clinical factors was explored for each cerebral compartment (white matter, grey matter, and cerebellum) and for the whole brain using a logistic regression analysis. A p-value of < 0.05 was taken as statistically significant.

The relationship between volumes, Kidokoro injury severity, and clinical factors was explored for each corresponding cerebral compartment (white matter, grey matter, and cerebellum) and for the whole brain using a multivariate linear regression analysis, considering the post-menstrual age at TEA-MRI as a covariate. A p-value of < 0.05 was taken as statistically significant. Some trends for p < 0.1 are also reported.

## Results

### Descriptive results

A total of 110 very and extreme preterm infants underwent a TEA-MRI between 38 to 42 weeks+ 6 days of PMA (Mean (SD) PMA: 41.1 (0.73)), 70 of whom were born at Robert Debré Hospital, and 40 were secondarily transferred. Among them, 54% (59/110) were males. Almost half of them required over a day of mechanical invasive ventilation and prolonged parenteral nutrition, 46% (51/110) and 45% (49/110) respectively. The mean (SD) GA at birth was 27.2 (2) weeks of gestation, with 30% (32/110) born before 26 weeks, 35 % (38/110) between 26 and 28 weeks, and 35% (38/110) after 28 weeks (**Table 1**).

**Table 1:** Description of the clinical population Abbreviations: PMA: postmenstrual age; MRI: magnetic resonance imaging; TEA: term equivalent age; SGA: Small for gestational age; BPD: Bronchopulmonary dysplasia

|  | n (%) |  |  | Mean (SD) |
| --- | --- | --- | --- | --- |
| **Males/Females** | 59 (54) / 51 (46) |  |  | - |
| **Gestational age at birth (weeks)** | **<26**<br>32 (30) | **26-28**<br>38 (35) | **>28**<br>38 (35) | 27.5 (2) |
| **Monochorionic twins** | 12 (11) |  |  | - |
| **SGA (<10<sup>th</sup> percentile)** | 24 (22) |  |  | - |
| **BPD** | 76 (69) |  |  | - |
| **Invasive ventilation > 1 day** | 51 (46) |  |  | - |
| **NEC** | 15 (14) |  |  | - |
| **Parenteral nutrition ≥ 21 days** | 49 (45) |  |  | - |
| **Sepsis** | 68 (62) |  |  | - |
| **PMA at TEA-MRI (weeks)** | - |  |  | 41.1 (0,73) |

The evaluation of preterm brains on TEA-MRI was first performed according to the Kidokoro scoring system. The intra-observer concordance coefficient Kappa for the whole brain score, based on repeated evaluations of 20 subjects, was 0.96 (95% CI: 0.90 – 1). Moderate to severe brain injuries were found in only 7% (7/110) of children, while most were classified as normal (70%) (77/110) (**Table 2**). Anomalies were more frequent in GM and cerebellum.

**Table 2:** Distribution of cerebral injury severity according to Kidokoro scoring, Hemorrhagic lesions distribution (in percentage of the 35% infants showing lesions) and DHESI frequency. * According to Papile classification. SEH: Subependymal hemorrhage, IVH: Intraventricular hemorrhage (grades 2, 3, 4)

| Kidokoro scoring: Severity | Normal | Mild | Moderate | Severe |
| --- | --- | --- | --- | --- |
| Whole brain | 70 % | 23 % | 4 % | 3 % |
| White matter | 78 % | 15 % | 3 % | 4 % |
| Grey matter | 65 % | 15 % | 16 % | 4 % |
| Cerebellum | 60 % | 19 % | 17 % | 4 % |
|  | SEH | IVH 2 | IVH 3 | IVH 4 |
| Hemorrhagic lesions* (35%) | 33 % | 36 % | 18 % | 13 % |
| DEHSI | 13 % |  |  |  |

Hemorrhagic lesions were identified in 35% of subjects; significant hemorragic lesions were taken into account in the Kidokoro scoring either with the ventricular dilatation for grade 3 hemorrhages or as white matter signal anomaly in case of grade 4 hemorrhagic lesions.

DEHSI (Diffuse excessive high signal intensity) was found in 13% of them (**Table 2**). The quantification of brain compartment volumes revealed high variability across subjects **(Figure 1b)**.

### Clinical factors associated with injury severity according to Kidokoro scoring

Multiple logistic regression analyses showed significant associations between the degree of brain anomaly according to the Kidokoro scoring and clinical factors **(Figure 2)**.

**Figure 2:**
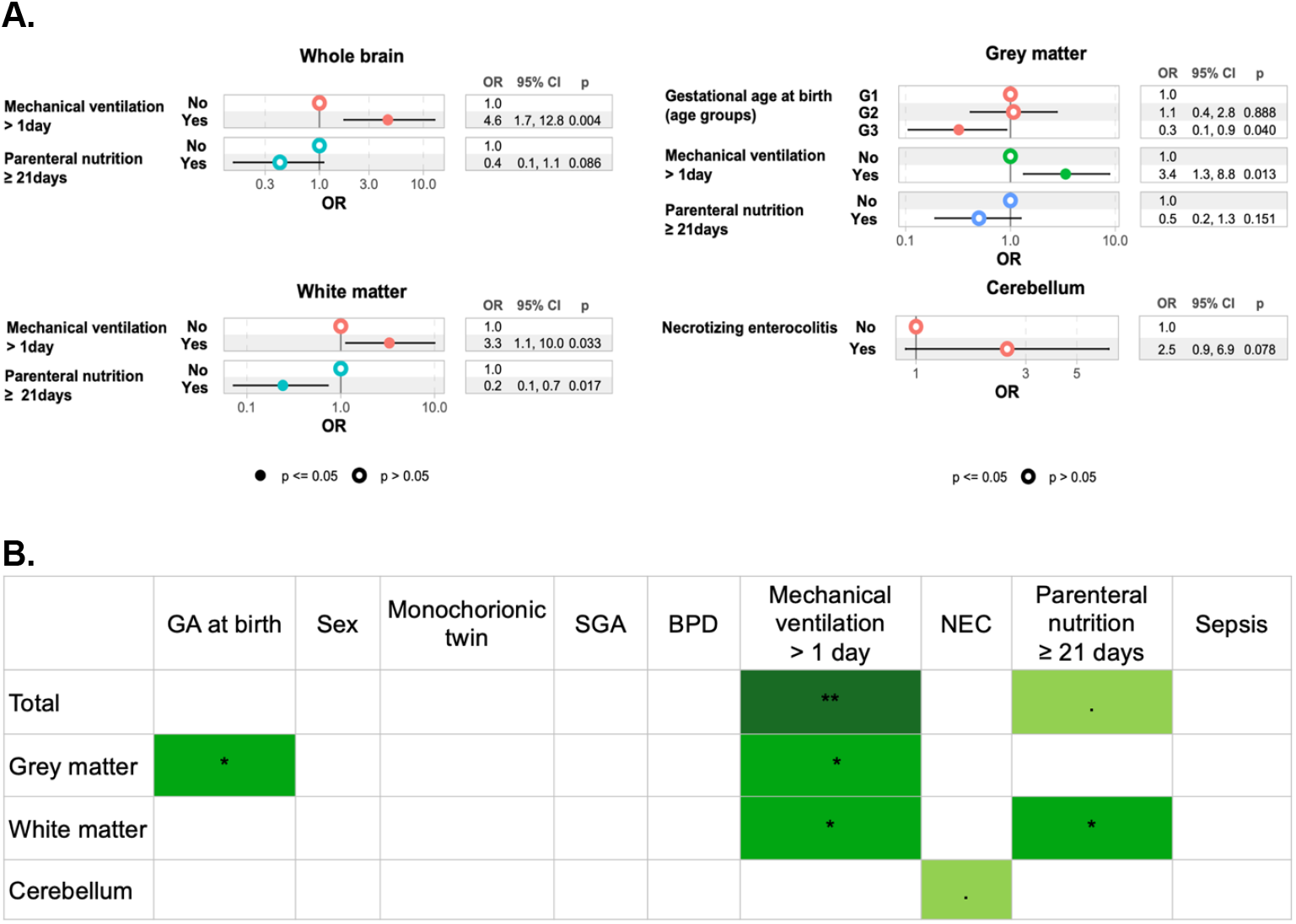
**A**. Associations between clinical factors and injury severity according to Kidokoro scoring, assessed with a logistic regression model (Odds-Ratio OR with their 95 % confidence interval CI) for the different brain compartments: **B**. Summary of the resulting significant associations and tendencies: **\*\*\*** p < 0.001 ; **\*\*** p < 0.01; **\*** p < 0.05; . p < 0.1 Abbreviations: GA: gestational age; G1: <26w GA, G2: 26-28w GA, G3: 28-32 GA; SGA: small for gestational age; BPD: bronchopulmonary dysplasia; NEC: necrotizing enterocolitis

In the whole brain, we showed that a higher degree of Kidokoro injury severity was significantly associated with mechanical invasive ventilation (adjusted Odds Ratio aOR = 4.6, 95% Confidence Interval CI [1.7 – 12.8], p = 0.004), whereas we observed a trend for a lower degree of severity for infants who underwent prolonged parenteral nutrition (aOR = 0.4, 95% CI [0.1-1.1], p= 0.086)

In grey matter, we found a similar association with mechanical invasive ventilation (aOR = 3.4, 95% CI [1.3 – 8.8], p = 0.013) but also with the category of gestational age at birth: the lower the gestational age at birth, the higher the severity (aOR = 0.3, 95% CI [0.1 – 0.9], p = 0.04).

In white matter, mechanical invasive ventilation was also identified as a risk factor for injury severity (aOR = 3.3, 95% CI [1.1 – 10], p = 0.033), and prolonged parenteral nutrition was identified as a protecting factor (aOR = 0.2, 95% CI [0.1 – 0.7], p = 0.017). No significant association was found between the clinical factors and injury severity in the cerebellum, yet children who had suffered from NEC had a tendency for higher injury severity (aOR = 2.5, 95% CI [0.9 – 6.9], p = 0.078).

### Clinical factors associated with cerebral volumes

Multiple linear regression was used to analyze cerebral volumes and their relationship with clinical factors, Kidokoro injury severity and PMA at TEA-MRI **(Figure 3)**.

**Figure 3:**
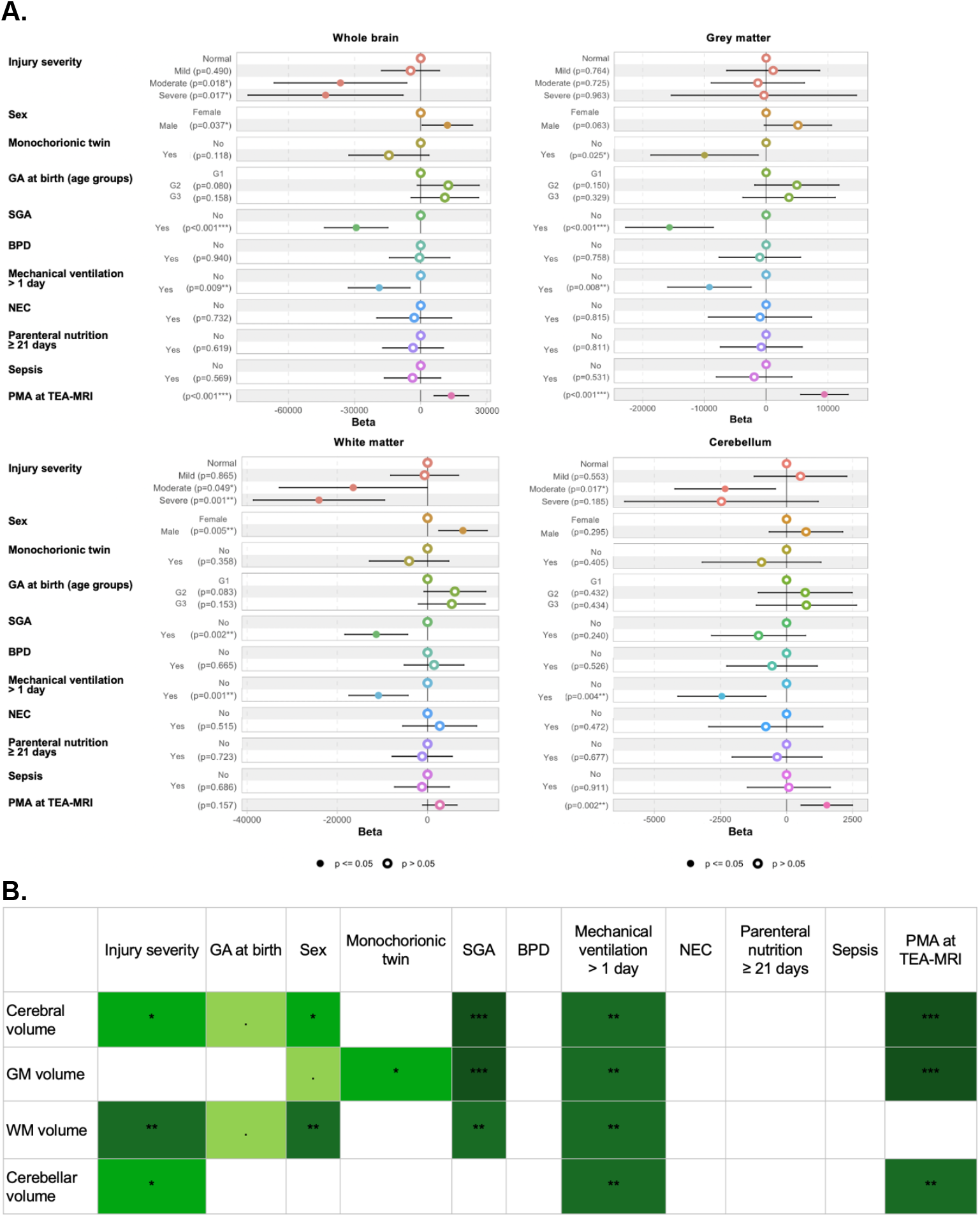
**A**. Associations between cerebral volumetry, injury severity according to Kidokoro scoring, and clinical factors assessed according to a linear regression model for the different brain compartments: the whole brain, grey matter, white matter, cerebellum. **B**. Summary of the resulting significant associations and tendencies: *** p < 0.001; ** p < 0.01; * p < 0.05; . p < 0.1 Abbreviations: GA: gestational age; G1: <26w GA, G2: 26-28w GA, G3: 28-32 GA; SGA: small for gestational age; BPD: bronchopulmonary dysplasia; NEC: necrotizing enterocolitis; PMA: postmenstrual age; MRI: magnetic resonance imaging; TEA: term equivalent age

First, volumes of the whole brain, GM and cerebellum showed a significant increase with PMA at TEA-MRI, justifying the need for controlling analyses for this variable.

The global cerebral volume was significantly reduced when moderate or severe lesions were present according to Kidokoro scoring (p = 0.018 and 0.017 respectively). Higher volumes were found in males (p = 0,037). Preterms born SGA (p < 0.001) or who had undergone mechanical invasive ventilation (p = 0.009) showed decreased volumes as well.

Grey matter volume was significantly reduced in children born as monochorionic twins (p = 0.025), SGA (p < 0.001), or having undergone mechanical invasive ventilation (p = 0.008). There was no association with brain injury severity according to Kidokoro scoring.

Concerning white matter volume, it was significantly reduced when moderate or severe lesions were present (p = 0.049 and 0.001 respectively), in children born SGA (p = 0.002) or having undergone mechanical invasive ventilation (p = 0.001). Males had higher white matter volumes (p = 0.005).

Concerning cerebellar volume, it was significantly reduced in children bearing moderate lesions (p = 0.017) or having undergone mechanical invasive ventilation (p = 0.004).

## Discussion

In this study, we found mechanical ventilation to be a strong risk factor for increased brain injury severity assessed with Kidokoro scoring in very and extreme preterm babies. Smaller GA at birth was found to be a risk factor for injury in grey matter, while prolonged parenteral nutrition seemed to act as a protective factor on white matter. Mechanical ventilation and being SGA were also found to be strong risk factors for a reduction in cerebral volumes. There was a clear association between increased injury severity and reduced cerebral volumes.

The distribution of brain injury severity we observed according to Kidokoro scoring was highly different from the one described in the original study, with only 7% of our patients showing moderate to severe injuries (compared to 35 % in the original study) [15]. Yet, this former study was set more than 10 years ago, and its population consisted in extremely premature newborns only. More recent studies revealed a distribution similar to ours, like the one from Abiramalatha et al. in 2022 with only 7% of patients having mild to severe injuries [23]. Still, this difference with the original study in injury severity and prevalence might explain a decreased significance of our results on relationships with clinical factors.

The association between mechanical ventilation and increased injury severity as well as reduced cerebral volumes had already been acknowledged in previous studies, such as the one from Brouwer et al. in 2017 [24]. This effect might be mediated through the cerebral inflammatory response to ventilation-induced oxidative stress [25], and / or the vasoconstrictive and ischemic effects of hypocarbia in the brain [26]. Our study further emphasizes the impact of mechanical ventilation on the early stages of cerebral development, but on the other hand we should keep in mind that this association might also reflect the severe clinical state of patients requiring such invasive ventilation.

Similarly, being SGA at birth was associated with reduced cerebral volumes (except for the cerebellum). Many children born SGA suffered from vascular intra-uterine growth restriction (IUGR), which increases the risk of chronic ischemia in the brain and might therefore explain a volume reduction in those children. Still, in previous research this reduction could be observed in all SGA preterms whether or not they had suffered from IUGR, implying other underlying mechanisms for this association. Interestingly, it was also shown that this cerebral volume reduction endures during infancy and adolescence, and is associated with the occurrence of neurodevelopmental disorders [27] [28] [29]. Nevertheless, in this study we could not exclude that children born SGA exhibited reduced cerebral volumes simply because of a lower weight, and not because of the reflection of a cerebral lesion.

Of note, male sex was associated with greater white matter volumes at TEA in our study. Still, this is most probably the consequence of sexual dimorphism, with boys exhibiting bigger cerebral volumes the same way their birth weight is higher than that of girls [30] [31] [32]: this would explain why boys have bigger volumes despite female sex being a known protective factor in preterms.

Low GA at birth seemed to increase injury severity, at least in grey matter. This was also observed in the original Kidokoro study [15] and by Brouwer et al. in 2017 [24]: the earlier the preterm is exposed to a possibly aggressive extra-uterine environment, the more the cerebral developmental course will be impacted. Still, this might not be the only explanation, and a higher degree of anomaly may exist from birth (or even before) in children with lower gestational age: the only way to assess this hypothesis would be to collect early MRI data, before TEA, in those preterm babies. Yet the Kidokoro scoring system was not developed to assess such early MRI-scans.

No association was found between cerebral volumes, severity of anomalies and BPD, NEC, or sepsis, a surprising result considering that those factors are important mediators of inflammation that might lead to cerebral injury. BPD has been found to increase the risk of behavioral disorders and reduce IQ in preterms [33] [34], yet studies by Abiramalatha et al. [23] or Brouwer et al. [24] could not show an association with cerebral injury severity at TEA. This suggests that BPD impact might be mediated by anomalies that are not easily identified with MRI. Furthermore, although BPD and sepsis were identified as risk factors in previous studies [10], it is interesting to note that those pathologies were considered only in their most severe forms previously. Lee et al. did show reduced cerebral and white matter volumes in BPD infants, yet this association disappeared when only mild BPD were studied [35]. Balakrishan et al. [36] did observe that sepsis increased injury severity but they only considered culture-positive sepsis, while it has already been proven that culture-positive and culture-negative sepsis do not lead to similar neurodevelopmental outcomes. Finally, we considered all stage ≥ II NEC, perforated and non-perforated, while it is known that patients with perforated NEC have higher odds of NDD than patients with non-perforated NEC [37]. Therefore, the lack of association in our study may be due to our choice in considering all kinds of BPD, both perforated and non-perforated NEC, and culture-negative sepsis in addition to culture-positive ones.

A more surprising result was that prolonged parenteral nutrition seemed to act as a protective factor against cerebral injury (at least on white matter). Indeed, in previous studies a prolonged parenteral nutrition was associated with reduced biparietal perimeter in preterms and delayed neurodevelopment [38]. Yet those results are controversial, as an early parenteral nutrition was also found to be related to higher cranial perimeter in preterms [39]. We might suspect that the impact of parenteral nutrition vary depending on its composition or preparation technique: indeed, Costa et al. found an increase in cerebellar volumes when using a multicomponent lipid emulsion [40], and it was shown that the increased use of in-line filtration techniques for the preparation of parenteral nutrition could reduce their concentration in micro-particles and the associated inflammatory response [41]. The optimization of parenteral nutrition might be an interesting approach to limit cerebral injury in preterms and improve their neurodevelopmental prognosis.

In our research, we observed a higher number of risk factors and higher significance when cerebral volumes were analyzed rather than injury severity with the Kidokoro scoring. In the same way, the study by Nosaka et al. highlighted lower white matter volumes at TEA for preterms suffering from choriamniotitis compared with controls but no difference in Kidokoro scores [11]. Analytic tools providing quantitative metrics such as cerebral volumes are more sensitive for the detection of cerebral anomalies in preterms than qualitative ones like the Kidokoro scoring, and therefore are more accurate to provide reproducible markers of prognosis. On the other hand, those quantitative analytic tools are not yet used in everyday practice, qualitative ones are more accessible and still hold some prognostic value, as suggested by the association between decreased cerebral volumes and increased anomaly severity according to Kidokoro scoring.

Our study suffered from some limitations, like its retrospective nature, allowing only for restricted access to clinical data, and its monocentric nature limiting comparability. Still, the children were born in different hospitals, our population benefitted from a certain degree of diversity. Furthermore, having realized all MRI on the same scanner, we reduced the variability in our image quality. The main limitation in our study was actually the limited number of patients exhibiting moderate-to-severe cerebral injuries since this confers disproportionate weight to the clinical characteristics of these few subjects. Also, we could not find a proper index of clinical severity to use as a confounding factor in our multivariate analysis, therefore increasing the impact of factors such as mechanical ventilation.

Despite those limitations, the strength of our study is to offer both a qualitative and quantitative analysis of the preterm brain at TEA, relying on a standardized and optimized MRI protocol. The collected dataset on very and extreme preterms will include neurodevelopment monitoring in the coming years, allowing us to further relate early neuroimaging results with outcome. This will also be the basis for future interventional studies, in which the risk and protective clinical factors that we identified could be used as confounding factors.

## Conclusion

This study provided an evaluation of the impact of clinical risk factors on early brain development in very and extreme preterms, emphasizing the negative impact of mechanical ventilation during the critical perinatal period, and demonstrating a protective role of parenteral nutrition. It further highlighted the benefits of using both a qualitative and quantitative analysis when exploring the preterm brain with TEA-MRI.

## Data Availability

Derived data generated will be shared on reasonable request to the corresponding author.

## Abbreviations

ADC: apparent diffusion coefficient
BPD: broncho-pulmonary dysplasia
CI: Confidence interval
DEHSI: diffuse excessive high signal intensity
FOV: field of view
GA: gestational age
GM: grey matter
MRI: magnetic resonance imaging
NDD: neurodevelopmental disorders
NEC: necrotizing enterocolitis
NICU: neonatal intensive care unit
OR: Odds-ratio
PMA: post-menstrual age
SGA: small for gestational age
WM: white matter

